# Natural Language Processing for Clinical Laboratory Data Repository Systems: Implementation and Evaluation for Respiratory Viruses

**DOI:** 10.1101/2022.11.28.22282767

**Authors:** Elham Dolatabadi, Branson Chen, Sarah A. Buchan, Alex Marchand-Austin, Mahmoud Azimaee, Allison J. McGeer, Samira Mubareka, Jeffrey C. Kwong

## Abstract

**Background:** With the growing volume and complexity of laboratory repositories, it has become tedious to parse unstructured data into structured and tabulated formats for secondary uses such as decision support, quality assurance, and outcome analysis. However, advances in Natural Language Processing (NLP) approaches have enabled efficient and automated extraction of clinically meaningful medical concepts from unstructured reports.

**Objective:** In this study, we aimed to determine the feasibility of using the NLP model for information extraction as an alternative approach to a time-consuming and operationally resource-intensive handcrafted rule-based tool. Therefore, we sought to develop and evaluate a deep learning-based NLP model to derive knowledge and extract information from text-based laboratory reports sourced from a provincial laboratory repository system.

**Methods:** The NLP model, a hierarchical multi-label classifier, was trained on a corpus of laboratory reports covering testing for 14 different respiratory viruses and viral subtypes. The corpus included 85*k* unique laboratory reports annotated by eight Subject Matter Experts (SME). The model’s performance stability and variation were analyzed across fine-grained and coarse-grained classes. Moreover, the model’s generalizability was also evaluated internally and externally on various test sets.

**Results:** The NLP model was trained several times with random initialization on the development corpus, and the results of the top ten best-performing models are presented in this paper. Overall, the NLP model performed well on internal, out-of-time (pre-COVID-19), and external (different laboratories) test sets with micro-averaged F1 scores >94% across all classes. Higher Precision and Recall scores with less variability were observed for the internal and pre-COVID-19 test sets. As expected, the model’s performance varied across categories and virus types due to the imbalanced nature of the corpus and sample sizes per class. There were intrinsically fewer classes of viruses being *detected* than those *tested*; therefore, the model’s performance (lowest F1-score of 57%) was noticeably lower in the “*detected*” cases.

**Conclusions:** We demonstrated that deep learning-based NLP models are promising solutions for information extraction from text-based laboratory reports. These approaches enable scalable, timely, and practical access to high-quality and encoded laboratory data if integrated into laboratory information system repositories.

## Introduction

Clinical laboratory data account for a large proportion of data stored in electronic health record systems worldwide and present a wealth of information vital for evidence-based decision-making and public health improvement ^1,2^. Laboratory information systems record, manage, and store laboratory test data to facilitate reporting to clinicians and jurisdictional laboratory information repositories ^3^. These repositories often include test orders and results from various laboratory service providers, such as hospitals, public health agencies, and private companies, and are populated as part of clinical care.

Several factors limit the secondary use of laboratory data for other purposes. The most important are concerns about the quality of the data, lack of standardization, and difficulty in extracting the needed information ^4,5^. Laboratory data varies over time due to evolving standards of care and changing population demographics. Furthermore, specific categories of laboratory data are reported as free text in an unstructured format with no standard vocabulary in the actual contents, which adds more complexity for their secondary uses ^1^. Therefore, efforts are needed to eliminate redundancies, extract the necessary information, and derive accurate interpretations from laboratory data.

Our institute has developed a specific information extraction workflow to manage the interpretation of a large volume of provincial clinical laboratory results, as shown in Figure 1. The workflow, called a semi-rule-based workflow, relies on time-consuming and operationally resource-intensive approaches, including a library of rule-based and hand-crafted tools. These tools are explicitly programmed for various laboratory result categories and must be refined continually. To address challenges with our existing semi-rule-based workflow and automate the exhaustive information retrieval task, we built a deep learning-based natural language processing (NLP) tool. The objective of this study was to assess the feasibility of our deep learning-based NLP model and evaluate its performance relative to the semi-rule-based workflow.

**Figure 1.**
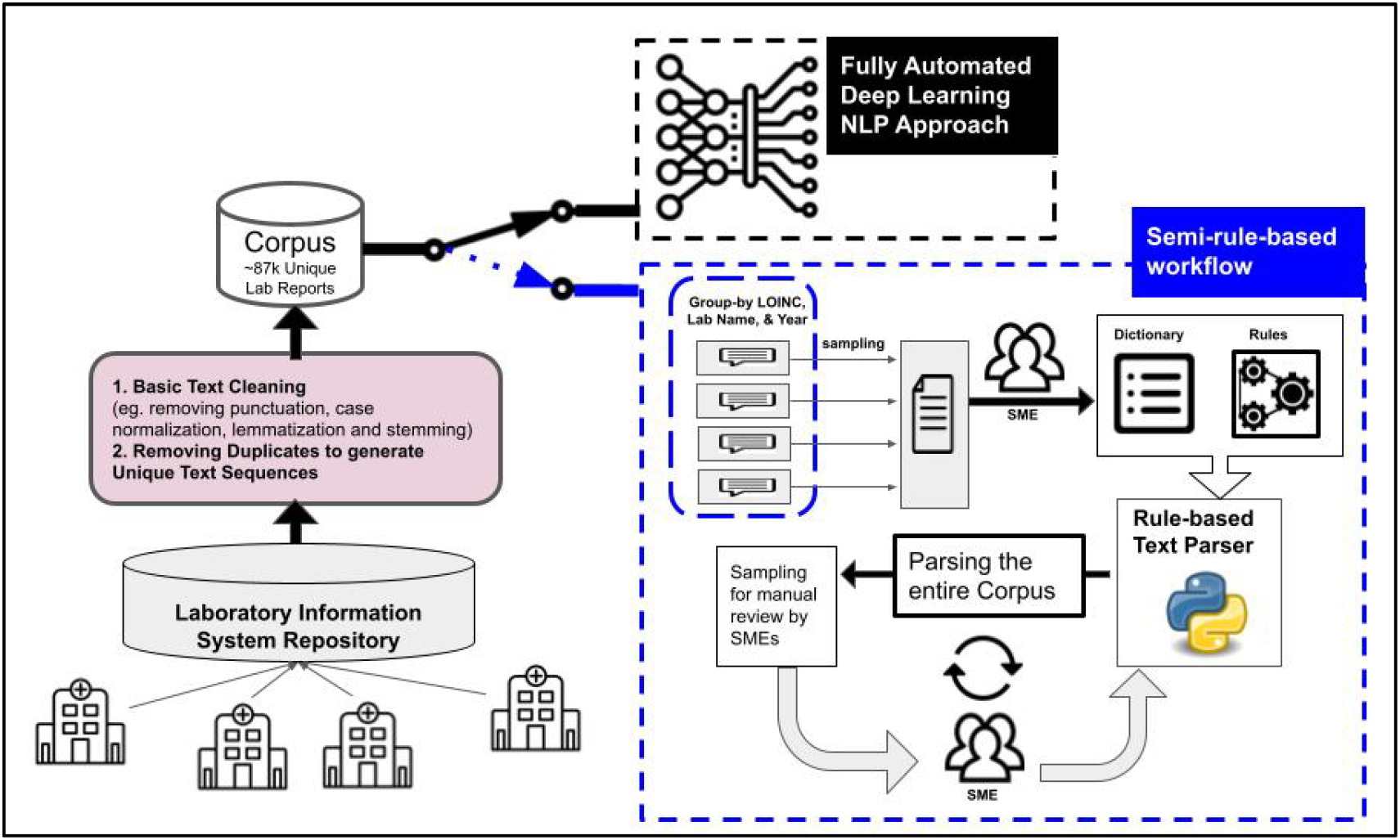
Semi-rule-based workflow vs. Fully automated Deep Learning NLP approach. Semi-rule-based relies on time-consuming and operationally resource-intensive approaches for the information extraction task. The corpus was derived from the Ontario Laboratories Information System (OLIS). OLIS has over 100 contributors, including hospital, commercial, and public health laboratories. Following basic text-cleaning steps, around 87k unique laboratory reports were collected and included in our corpus to be used in parallel by both approaches: Semi-rule-based and Deep Learning NLP. Semi-rule-based workflow is a multistep procedure where all the unique reports were grouped by the LOINCs, year, and location in the first step. In the second step, SMEs created a list of dictionaries for terms related to the different viruses and strains and a set of *if-then-else* rules to generate interpretations and extract information from each laboratory report. The dictionaries and if-then-else rules were packaged as a python library called the rule-based text parser. Finally, the parser was improved based on inputs from three SMEs in an iterative manner (more details in the Supplementary Materials).

The development of NLP methods is essential to automatically transform laboratory reports into a structured representation that scales data usability for research, quality improvement, and clinical purposes ^6–12^. NLP enables automated extraction of information, and its use in the clinical domain is growing with increasing uptake in various applications such as biomedical named entity recognition ^11,12^, summarization ^10^, and clinical prediction tasks ^9^. More recently, deep learning approaches such as convolutional neural networks (CNN), recurrent neural networks (RNN), and RNN variants such as bidirectional Long Short-Term Memory (Bi-LSTM) have been successfully applied to clinical NLP tasks ^10,13–16^. They are now considered the baseline techniques for various information extraction tasks ^11,12,17–20^.

In this study, we focused on automating the retrieval of information related to respiratory viruses from the laboratory repository of Canada’s most populous province. Respiratory viruses account for a substantial burden of disease globally ^21,22^, causing respiratory and non-respiratory illnesses ^23^. It is impossible to distinguish which respiratory virus is causing infection based on clinical examination alone, necessitating laboratory testing for confirmation. We sought to (1) implement a deep-learning-based NLP predictive model to extract respiratory virus information from the laboratory repository and (2) evaluate the generalizability and robustness of predictions (extracted information) across different categories of respiratory viruses and test sets. This study’s findings can inform public health practitioners and researchers about using NLP approaches to empower and facilitate access and retrieval of information from a collection of text-based laboratory reports without any previous time-consuming handcrafted rule-based approaches. Furthermore, it leads to a scalable and easily deployable automated information extraction tool.

## Methods

### Study Design

The dataset used in this study is a collection of laboratory reports that cover testing for 14 different respiratory viruses and viral subtypes (Table 1), most of which are in the form of texts. The reports are text-based and require cleaning, parsing, and encoding.

**Table 1:**
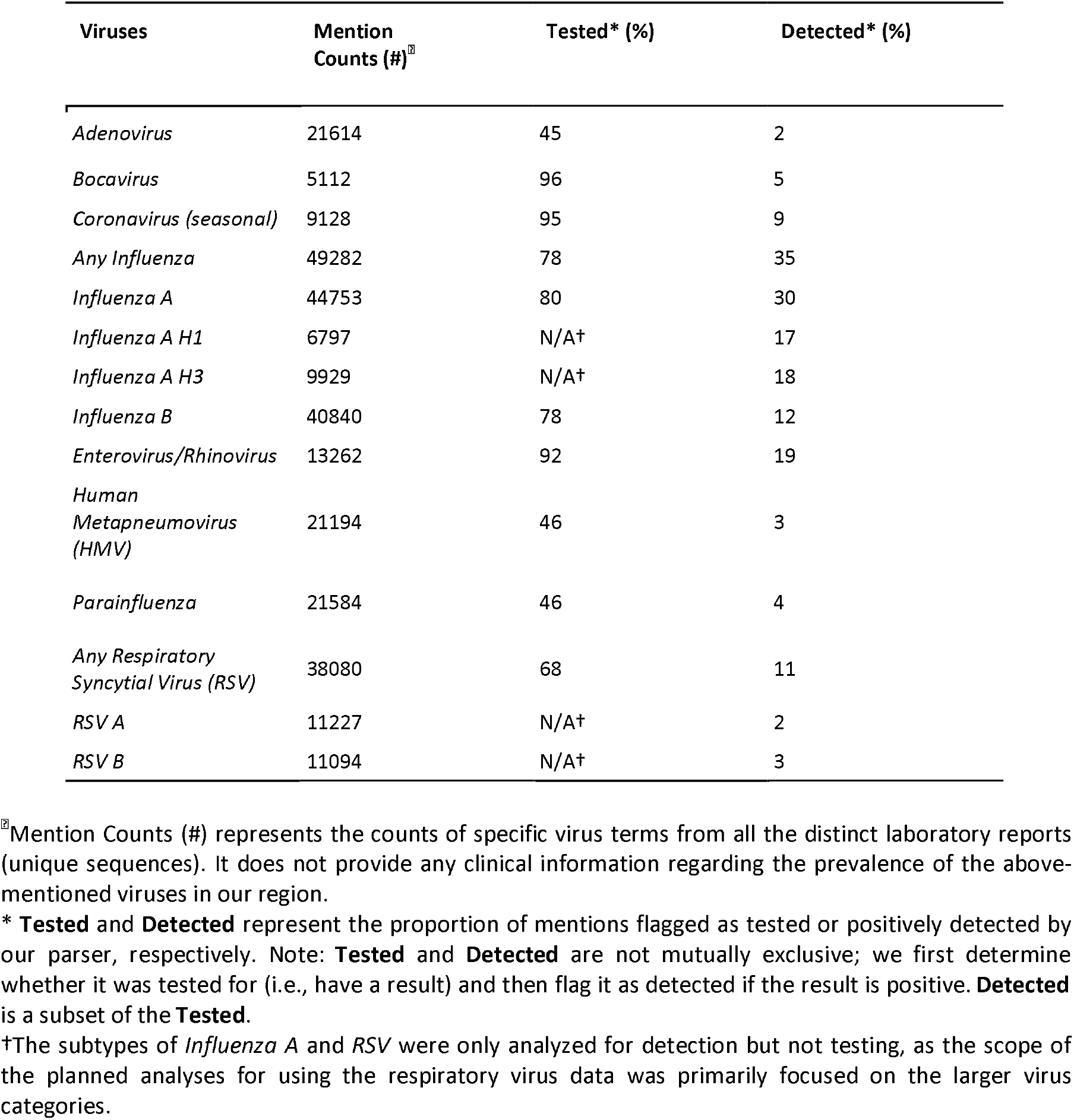
Details of the respiratory viruses embedded in text-based laboratory reports derived from the Ontario Laboratories Information System (OLIS). Specimens may be tested for one or more of the following viruses: *influenza, respiratory syncytial virus (RSV), adenoviruses, seasonal coronaviruses, enterovirus/rhinoviruses, parainfluenza viruses, human metapneumovirus (HMV), and bocavirus*. The testing modalities employed include single and multiplex polymerase chain reaction, direct fluorescent antibody, viral culture, and enzyme immunoassay rapid antigen tests. Repeated testing may involve multiple laboratories and testing modalities.

| Viruses | Mention Counts (#)<sup>¶</sup> | Tested* (%) | Detected* (%) |
| --- | --- | --- | --- |
| *Adenovirus* | 21614 | 45 | 2 |
| *Bocavirus* | 5112 | 96 | 5 |
| *Coronavirus (seasonal)* | 9128 | 95 | 9 |
| *Any Influenza* | 49282 | 78 | 35 |
| *Influenza A* | 44753 | 80 | 30 |
| *Influenza A H1* | 6797 | N/A<sup>†</sup> | 17 |
| *Influenza A H3* | 9929 | N/A<sup>†</sup> | 18 |
| *Influenza B* | 40840 | 78 | 12 |
| *Enterovirus/Rhinovirus* | 13262 | 92 | 19 |
| *Human Metapneumovirus (HMPV)* | 21194 | 46 | 3 |
| *Parainfluenza* | 21584 | 46 | 4 |
| *Any Respiratory Syncytial Virus (RSV)* | 38080 | 68 | 11 |
| *RSV A* | 11227 | N/A<sup>†</sup> | 2 |
| *RSV B* | 11094 | N/A<sup>†</sup> | 3 |

<sup>¶</sup> Mention Counts (#) represents the counts of specific virus terms from all the distinct laboratory reports (unique sequences). It does not provide any clinical information regarding the prevalence of the above-mentioned viruses in our region.
\* **Tested** and **Detected** represent the proportion of mentions flagged as tested or positively detected by our parser, respectively. Note: **Tested** and **Detected** are not mutually exclusive; we first determine whether it was tested for (i.e., have a result) and then flag it as detected if the result is positive. **Detected** is a subset of the **Tested**.
<sup>†</sup>The subtypes of *Influenza A* and *RSV* were only analyzed for detection but not testing, as the scope of the planned analyses for using the respiratory virus data was primarily focused on the larger virus categories.

The dataset was derived from the Ontario Laboratories Information System (OLIS). OLIS has over 100 contributors, which comprise hospital, commercial, and public health laboratories, adding to the complexity and variability of the clinical data. These data were analyzed at ICES (formerly the Institute for Clinical Evaluative Sciences).

The automated encoding of laboratory testing reports into respiratory viruses is framed as a multi-label classification task according to a hierarchy. There are two levels of classification hierarchy; at each level, the classification is multi-label. Each input text sequence can be assigned to a non-empty subset of various labels, as shown in Figure 2. In the first level of the hierarchy, the classifier assigns outputs to mutually non-exclusive fine-grained labels. The fine-grained labels are reassigned to a coarse-grained set of labels in the second level of the classification hierarchy. In this work, “sequence” refers to the input laboratory reports to the NLP model, which may be a single or several sentences. A “label” also refers to a status of a specific virus or strain being tested or ***Detected*** (e.g., *influenza A is detected*). ***Tested*** and ***Detected*** are not mutually exclusive; it must first be determined if the specimen is tested for any virus and then flagged as detected if the result is positive. ***Detected*** is a subset of the ***Tested***.

**Figure 2.**
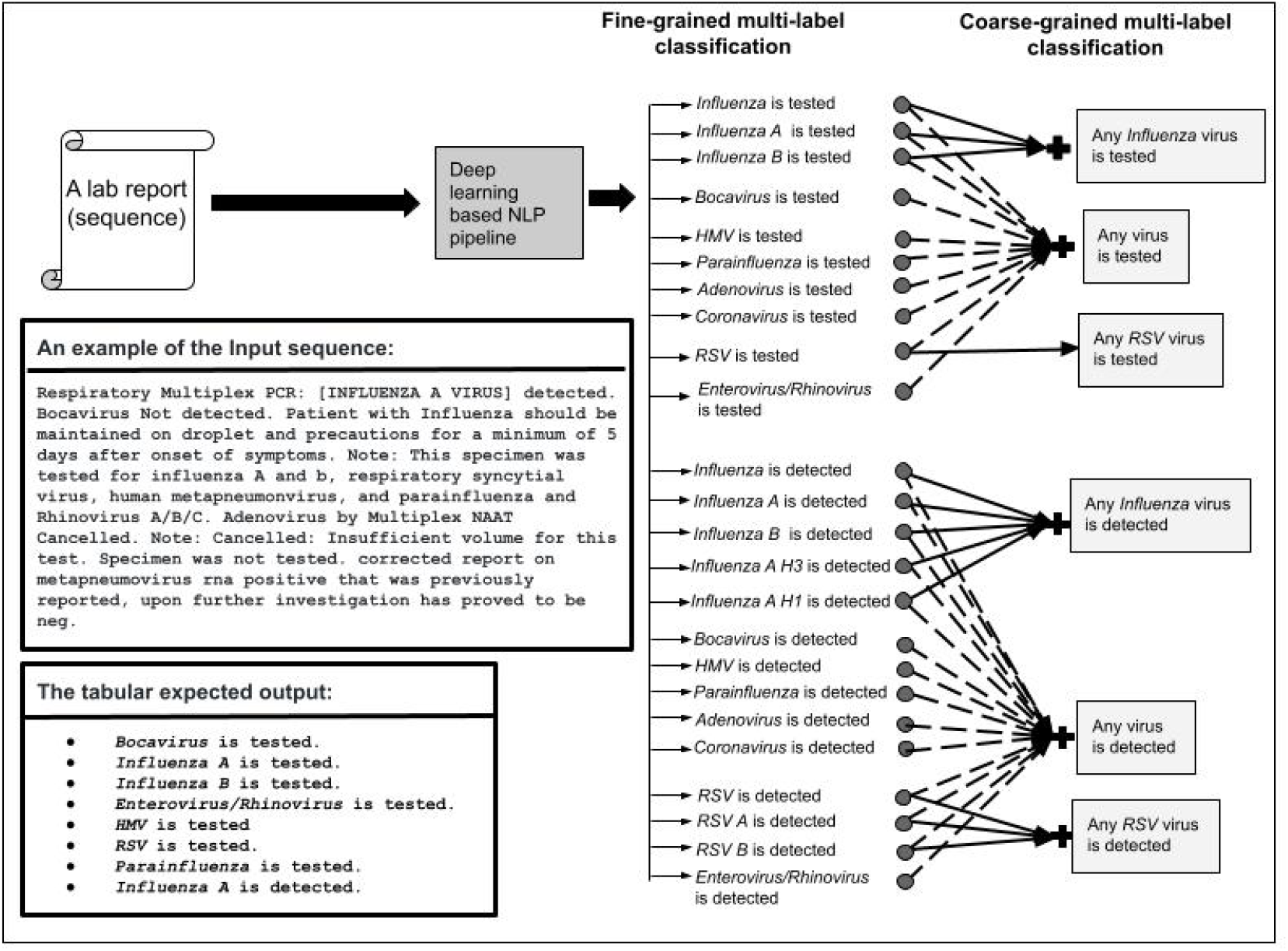
The fully automated deep learning-based NLP approach is a hierarchical-based multilabel classification task that retrieves virus (or strain) types and identifies their status as being tested and/or detected. Each input sequence was assigned to 24 mutually non-exclusive fine-grained labels in the first level of the hierarchy. In the second level of the classification hierarchy, the fine-grained labels were reassigned to a coarse-grained set of labels (k=6). An example laboratory report processed through the deep learning-based NLP approach for automated extraction and encoding of information is also shown. Notes: A *sequence* refers to the input laboratory reports to the NLP approach, which may be a single or several sentences. A *label* also refers to a status of a specific virus or strain (tested or detected). *Influenza is tested* implies it was tested for any *Influenza* type; however, the total number of ‘*Influenza is tested*’ is greater than the total number of *Influenza A tested + Influenza B tested* since not all *Influenza* is mentioned for types. The same applies to “*Influenza is detected*” and “*RSV is tested*”.

To summarize, the information extraction for an input text sequence involves retrieving virus types and identifying their status as being tested and(or) detected. Figure 2 illustrates a running example of the input and output of the deep learning-based NLP model.

### Corpus Development Description

#### Ontario Laboratories Information System (OLIS)

To create the corpus for this study, over a million observations corresponding to 99 unique Logical Observation Identifiers Names and Codes (LOINCs) were pulled from OLIS, and the text-based laboratory results were extracted from the observations. OLIS was created and is managed by Ontario Health, from whom ICES receives an ongoing data feed. At the time of writing, the OLIS data holding at ICES consists of >9,000 unique LOINCs and >5 billion laboratory observations across 150 laboratory test centers in Ontario. As such, the clinical laboratory data has considerable complexity and variability.

#### Development of the Ground Truth

In this study, we leveraged the semi-rule-based workflow, an information extraction workflow relying on a library of rule-based and hand-crafted tools, to create ground truth for the deep learning model. A group of eight Subject Matter Experts (SME), including two infectious disease epidemiologists, two infectious disease microbiologists, a genomic specialist, a research methodologist, a data analyst, and a machine learning scientist, were engaged in performing the required tasks in the workflow. These tasks include basic text cleaning, quality checking, and rule-based algorithm development for interpreting reports, as shown in Figure 1. In our institute, LOINCs are mainly used to filter OLIS observations into relevant groupings (e.g., respiratory viruses) and not for encoding and interpretation since they are not always used appropriately by those entering the data into OLIS. Consequently, the SMEs identified a list of 99 LOINCs related to respiratory viruses, and all the laboratory reports in OLIS corresponding to these LOINC codes were retrieved. The workflow consists of the following tasks:

1. The data analyst and data scientist first scanned the text strings and, after performing basic text cleaning (e.g., removing punctuations, stop words, case normalization, lemmatization, and stemming) and removing duplicates, they created a meaningful list of 87k unique laboratory reports.
2. The unique reports were grouped by laboratory and facility names, LOINCs, and year. Then, three SMEs, including two analysts and an infectious disease specialist, manually reviewed multiple samples per group and created a knowledge base and sets of *if-then-else* rules to generate interpretations for each laboratory report. Specifically, the knowledge base consisted of dictionaries for terms related to the different viruses and strains. The *if-then-else* rules provided instructions for grouping virus terms with respective results packaged as a python library, which we refer to in this study as the rule-based text parser.
3. Following the initial development of the rule-based text parser, it was improved based on inputs from three other SMEs in an iterative manner. The text parser was applied to the entire corpus to generate annotations at each iteration. Then the data analyst manually reviewed the interpretations and flagged unclear results to be reviewed by SMEs at another iteration. In addition, a small random sample of unflagged test results was provided to SMEs to be reviewed at this iteration. The SMEs subsequently reviewed the list and provided new rules to be added to the text parser. This procedure was repeated until there were no more flagged test results.

### Model Development and Evaluation

#### NLP Model Description

The deep learning-based NLP model consisted of three components that were trained jointly: the Word Embedding layer, the Bidirectional long short-term memory (Bi-LSTM) layer, and the Output layer. The word embedding layer computed a vector representation of each word in the text as a combination of a character-based representation learning model ^24^,^25^, and word vectors initialized with pre-trained GloVe embedding ^26^. The embedding layer was coupled with a Bi-LSTM on top of it to generate conceptually and contextually meaningful representations of words. An output layer of size equal to the number of distinct labels was placed on top of Bi-LSTM, and the last hidden state of the BiLSTM was projected into the output layer.

#### Model Evaluation

The model’s robustness and generalizability were evaluated internally and externally on various test sets, as in Table 2. The *internal* test set used for model training was a randomly sampled subset representing 10% of the laboratory reports from OLIS from 2007 to 2018. The performance of the model was also evaluated on two *out-of-time* test sets, including samples from an entirely different time period: (1) a large *pre-COVID-19* (2019) sample and (2) a small *post-COVID-19* (2020) sample. A separate test set, denoted as the *external* test set, included samples up to 2019 from two separate laboratories (testing sites not included in the development of the model) and was used to assess the external generalizability of the model. F1, Precision, and Recall scores were calculated for the model’s predictions. The paired t-test was used to determine whether a statistically significant difference in the F1 scores between classes and test sets existed. In addition, 95% confidence intervals were calculated for the Precision and Recall scores to quantify the uncertainty of the model’s estimates.

**Table 2.** Dataset statistics for laboratory descriptions of the development and test sets.

| Cohorts |  | seq(#)<sup>□</sup> | Any Influenza virus* |  | Any RSV virus |  | Any virus |  |
| --- | --- | --- | --- | --- | --- | --- | --- | --- |
|  |  |  | detected (#) | tested (#) | detected (#) | tested (#) | detected (#) | tested (#) |
| Development set<br>(years 2009-2018) | Training set | 60471 | 13792 | 35292 | 3959 | 27196 | 22284 | 40652 |
|  | Internal test set | 6719 | 1604 | 3941 | 428 | 3009 | 2541 | 4534 |
| Out-of-time test sets | pre-COVID-19 (2019) | 15908 | 3019 | 6903 | 706 | 5957 | 4745 | 8643 |
|  | post-COVID-19 (2020) | 100 | n/a | 11 | <6 | 11 | <6 | 27 |
| External test set<br>(years 2009-2018) |  | 4213 | 864 | 3020 | 261 | 2546 | 1431 | 3237 |

## Results

The development corpus, including training and test sets, included 87,500 sequences involving ∼5 million tokens (summary statistics for the datasets are shown in Table 2). The NLP model was implemented in Tensorflow on an NVidia Tesla GPU, and Adam was used as the optimization algorithm. The maximum sequence length was fixed to 400 words. The model was trained several times with random initialization on the development corpus, and the results of the top ten best-performing models on the test sets are presented here. The results for the fine-grained classification in the first level of the hierarchy are aggregated by micro-averaging across fine-grained 24 labels and are shown in Table 3. The detailed performance for each label is also shown in Supplementary Table 1. The F1-score performance of the model in the second level of the hierarchy, coarse-grained multi-label classification, for ‘*Any Influenza*’, ‘*Any RSV*’, and ‘*Any Virus*’ are also shown in Table 3. In addition, the variation of the model’s Precision and Recall scores using barplots and 95% confidence intervals are shown in Figure 3.

**Table 3.**
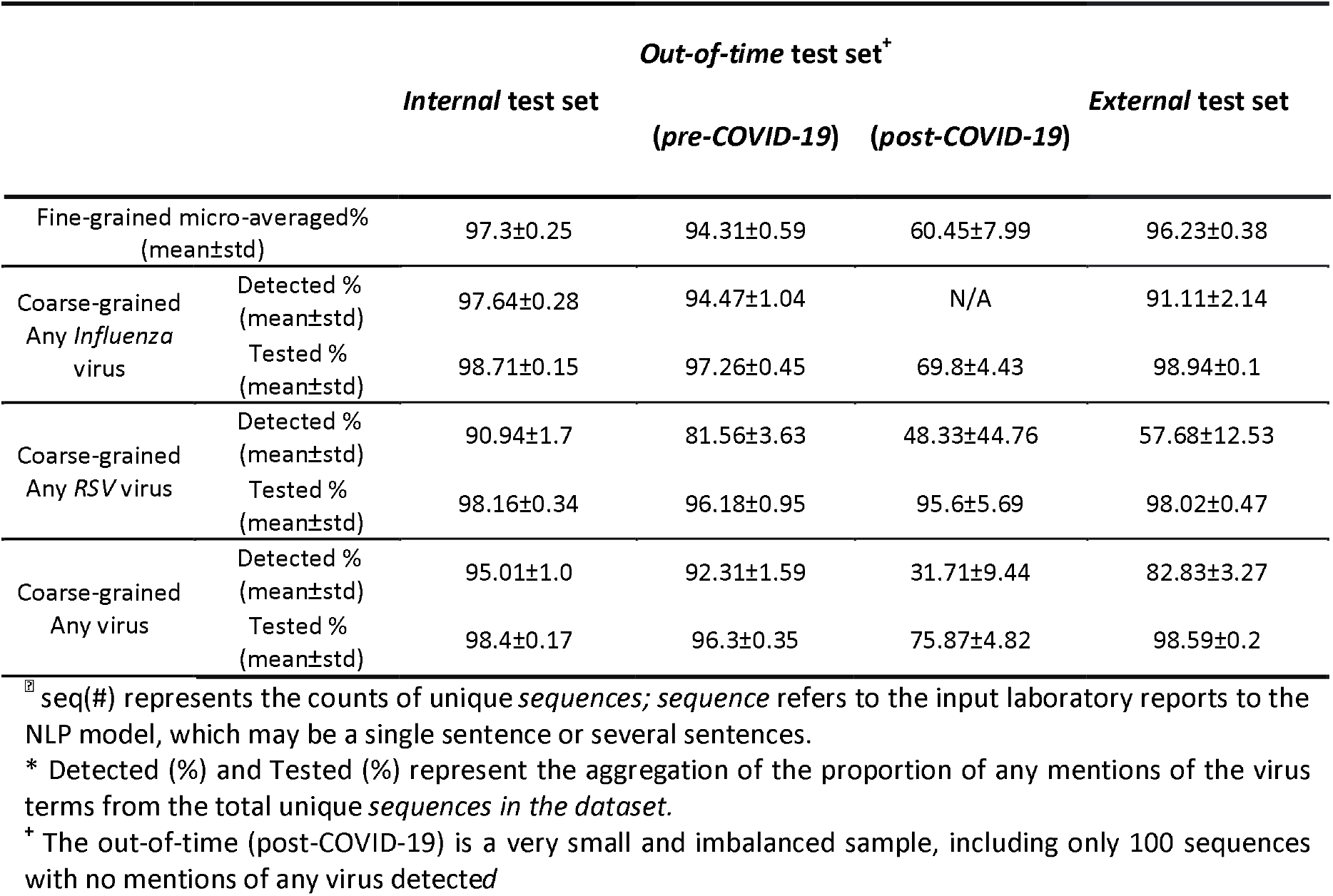
The prediction results (F1-score) of the top 10 best-performing models on the *in-time, out-of-time*, and *external test* sets. The fine-grained results are aggregated by micro-averaging across 24 fine-grained labels. The fine-grained results with the best-performing model are shown in Supplementary Table 1.

**Figure 3.**
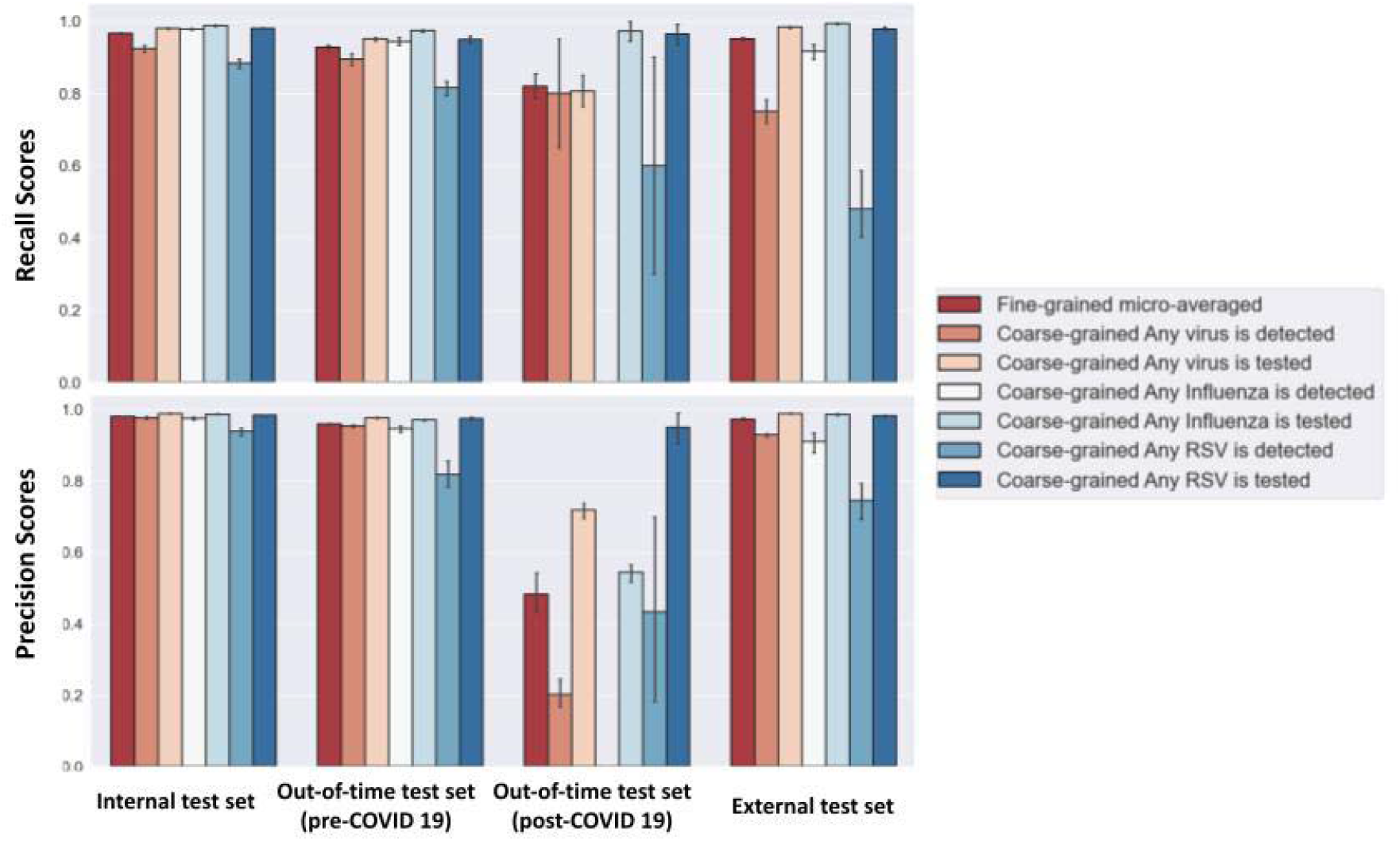
The Precision and Recall scores of the predictions of the top 10 best-performing models with 95% confidence intervals. The fine-grained results are aggregated by micro-averaging across 24 fine-grained labels.

As expected, the performance on the internal test set was better than the *out-of-time* (*pre-COVID-19*) and *external* test sets. In this regard, the F1 score results of the test sets were compared, and noticeable differences were observed between the pairs of *internal* and *out-of-time* (*pre-COVID-19*) test sets, *internal* and *out-of-time* (*post-COVID-19*) test sets, and *internal* and *external* test sets. The out-of-time (*post-COVID-19*) test set was a small and imbalanced sample, including 100 sequences with <6 mentions of any virus as being detected. The sample included 12 sequences labelled as being tested for coronavirus, and our model correctly classified them with an F1 score of 0.67. Regarding the degree of uncertainty in the estimates, fewer variations in Precision and Recall scores are observed for the *internal* and *out-of-time* test sets (*Pre-COVID-19*). On the contrary, the estimates on the *out-of-time* (*post-COVID-19*) and *external* test sets have larger confidence intervals.

In general, the models’ estimates on any test sets were variable across classes with varying degrees of uncertainty. The averaged F1 scores of the estimates for both “fine-grained (micro-averaged)” and “coarse-grained Any virus” classes were above 90% on the internal test set. The F1 score for the coarse-grained ‘*Any Influenza detected*’ on all test sets was above 91%. Overall, the performance for “*Coarse-grained detected*” classes was lower than for “*Coarse-grained tested*” classes. Among the detected classes, the performance for ‘*Any Influenza virus*’ was evidently higher than ‘*Any RSV virus’*. The same result was observed between ‘*Any Influenza virus*’ and ‘*Any RSV virus’*. Comparably, larger confidence intervals are evidenced for the ‘*Coarse-grained Any RSV detected’* estimates.

## Discussion

### Principal Results

In this study, we demonstrated an implementation and evaluation of an NLP model for an automated and reductive information extraction task in a province-wide laboratory data repository. Our results suggest that the NLP model is a promising approach for information extraction from text-based laboratory reports as an alternative method to address the time-consuming and operationally resource-intensive nature of handcrafted rule-based models.

#### Model’s Generalization across Various Test Sets

Overall, the NLP solution, which was a hierarchical multi-label classifier, performed well on *internal, out-of-time* (*pre-COVID-19*), and *external* (different laboratories) test sets; except for the internal test sets, the other test sets were sourced from either a more recent time period or other laboratory sites, but the model was able to generalize well with micro-averaged F1 score >94% across all classes. The performance of the model on the other out-of-time (post-COVID-19) test set was satisfactory, however, due to its small sample size with many underrepresented classes, it was not possible to draw any conclusion. The out-of-time (post-COVID-19) test set was pulled from the 2020 cohort to simulate a non-stationary production environment for observation.

#### Models’ Stability and Performance Variation between Classes

In general, the model performances on any test sets were variable across classes and virus types due to the imbalanced nature of the corpus and sample sizes per class. There were intrinsically fewer classes of viruses *detected* compared with those *tested*. Therefore, the model’s performance was noticeably lower in the “*detected*” cases. Among *detected* cases, the lowest performance was observed for *RSV A*, and the highest performance among the *tested* cases was observed for *Influenza*. Moreover, more considerable variations were observed for the positive predictive and sensitivity values of the “*detected*” classes and, in particular, for the “*Any RSV virus detected*” class.

### Limitations

Although deep learning models promise great potential for digitized health data, putting the model into production and prospectively validating operational data is as crucial as model building and are a critical step in assessing and ensuring its operational effectiveness. However, we expect the model’s performance to deteriorate as it goes into production, potentially impacting data quality. Moving forward, we plan to run a silent-period production validation to further prospectively explore the model’s performance. During the silent period, our model will be integrated into the data quality and management workflow for the laboratory data repository, and the outputs will be internally validated in a fashion that would avoid exposure to data users. We also plan to run rigorous evaluation and continuous refinement of the model in the silent period to assess its performance better before it enters production.

Another significant limitation of this study is that the model was only trained on respiratory virus laboratory reports. Even within that collection, some categories were naturally underrepresented, which impacted the model’s generalizability. So, during the silent period, more records from a diverse set of laboratory reports from various categories will be annotated and made available to the model, and the model will be updated accordingly.

### Comparison with Prior Work

Deep learning-based NLP approaches have shown their efficiency in many clinical NLP tasks and have thoroughly permeated the informatics community. The existing body of literature has mainly focused on using deep learning models to extract and interpret cancer-related clinical concepts ^17,27,28^ from free text or other clinically meaningful entities from radiology reports or hospital notes ^10,15^. Only one study at the time of writing explored the use of an NLP system, Topaz, for the automated extraction and classification of influenza-related terms from text emergency reports ^29–31^. To our knowledge, our study is the first to explore using deep learning models for efficient processing and extraction of clinically meaningful knowledge pertaining to respiratory viruses from a laboratory repository.

One strength of the NLP approach used in this study is its scalability for various text-based laboratory scenarios. As the size and complexity of laboratory data grow, so does the need for scalable and reusable tools for automated extraction of knowledge from vast amounts of clinical notes and quick generalization from one task to another. Manual processing of laboratory reports severely limits the utilization of rich information embedded in the data repositories and makes the process of data cleaning and quality improvement prohibitively expensive. Usually, the rules learned from cleaning one collection of laboratory reports show little generalizability toward other collections. On the other hand, deep learning-based NLP algorithms are well-poised to scale the information extraction process. Although building deep learning-based NLP models is computationally intensive and memory demanding, the benefit-to-cost ratio of these models in clinical settings will continue to increase.

## Conclusions

The health industry is rapidly becoming digitized, and information extraction is a promising method for researchers and clinicians seeking quick retrieval of information embedded in texts. This study described developing and validating a deep learning-based NLP approach to extract respiratory virus testing information from laboratory reports. We demonstrated that our system can classify and encode large volumes of text-based laboratory reports with high performance without any of the previous time-consuming handcrafted feature engineering approaches. Taken together, the findings in this study provide encouraging support that NLP-based information extraction should become an important component of laboratory information repositories to assist researchers, clinicians, and healthcare providers with their information and knowledge management tasks.

## Data Availability

The data underlying this work is held securely in the coded form at ICES and, therefore, cannot be shared publicly due to data privacy concerns and legal data sharing agreements between ICES and data providers (e.g., healthcare organizations and government). However, data access might be granted to those who meet prespecified criteria for confidential access, available at www.ices.on.ca/DAS.

https://www.ices.on.ca/Data-and-Privacy/ICES-data

## Acknowledgements

This study was a collaborative effort supported by the Vector Institute, an independent, not-for-profit corporation dedicated to research in the field of artificial intelligence (AI), and ICES, an independent, non-profit research organization that uses population-based health and social data to produce knowledge on a broad range of health care issues. Resources used in preparing this work were funded by an annual grant from the Ontario Ministry of Health (MOH) and the Ministry of Long-Term Care (MLTC). Parts of this material are based on data and information compiled and provided by the Ontario Ministry of Health. This work was also supported by a SickKids-Canadian Institutes of Health Research New Investigator Grant in Child and Youth Health (NI19-1065). The analyses, conclusions, opinions, and statements expressed herein are solely those of the authors and do not reflect those of the funding or data sources; no endorsement is intended or should be inferred.

## Conflicts of Interest

The authors have no competing interests to declare.

### Abbreviations

NLP: Natural Language Processing
LIS: Laboratory Information System
OLIS: Ontario Laboratory Information System
Bi-LSTM: bidirectional Long Short-Term Memory
EHR: Electronic Health Record
RSV: Respiratory syncytial virus
HMV: Human Metapneumovirus
SME: subject matter experts
CI: Confidence Interval

## ETHICS STATEMENT

Projects that use data collected by ICES under section 45 of Ontario’s Personal Health Information Protection Act (PHIPA), and use no other data, are exempt from research ethics board review. The use of the data in this project is authorized under section 45 and approved by ICES’ Privacy and Legal Office. ICES is a prescribed entity under PHIPA. Section 45 of PHIPA authorizes ICES to collect personal health information, without consent, for the purpose of analysis or compiling statistical information concerning the management of, evaluation or monitoring of, the allocation of resources to or planning for all or part of the health system.

