## Supplemental Table 1 for "Natural Language Processing for Clinical Laboratory Data Repository Systems: Implementation and Evaluation for Respiratory Viruses"

**Supplementary Table 1** Fine-grained classification results (F1-Scores (%) from the best performing model).

| Labels | In-time test set | Out-of-time test set (Pre-COVID19) | Out-of-time test set (Post-COVID19) | External test set |
|---|---|---|---|---|
| Adenovirus is detected | 65.65 | 80.76 | 0.00 | 47.89 |
| Adenovirus is tested | 97.49 | 96.78 | 88.89 | 98.11 |
| Bocavirus is detected | 62.07 | 14.29 | 0.00 | 0.00 |
| Bocavirus is tested | 97.95 | 71.97 | 0.00 | 97.75 |
| Coronavirus (seasonal) is detected | 72.57 | 74.55 | 0.00 | 27.27 |
| Coronavirus is tested | 97.99 | 92.73 | 66.67 | 96.75 |
| Any Influenza is detected | 97.78 | 95.38 | 0.00 | 94.45 |
| Any Influenza is tested | 98.73 | 97.64 | 64.52 | 98.92 |
| Influenza A is detected | 97.50 | 95.68 | 0.00 | 89.47 |
| Influenza A is tested | 98.33 | 97.23 | 50.00 | 98.91 |
| Influenza B is detected | 94.64 | 68.91 | 0.00 | 53.91 |
| Influenza B is tested | 98.54 | 97.19 | 45.45 | 99.02 |
| Influenza A H1 is detected | 85.53 | 74.82 | 0.00 | 75.76 |
| Influenza A H3 is detected | 82.19 | 64.22 | 0.00 | 44.04 |
| Enterovirus/Rhinovirus is detected | 89.45 | 86.96 | 100.00 | 45.67 |
| Enterovirus/Rhinovirus is tested | 97.70 | 93.37 | 72.73 | 96.10 |
| Human Metapneumovirus (HMV) is detected | 85.44 | 78.49 | 0.00 | 60.47 |
| Human Metapneumovirus (HMV) is tested | 98.60 | 98.14 | 88.89 | 99.23 |
| Parainfluenza is detected | 90.08 | 83.20 | 0.00 | 84.30 |
| Parainfluenza is tested | 98.15 | 96.82 | 88.89 | 98.94 |
| Any Respiratory Syncytial Virus (RSV) is detected | 92.25 | 83.08 | 66.67 | 55.98 |
| Any Respiratory Syncytial Virus (RSV) is tested | 98.13 | 96.18 | 90.91 | 97.52 |
| RSV A is detected | 22.22 | 2.44 | 0.00 | 0.00 |
| RSV B is detected | 62.86 | 17.39 | 0.00 | 0.00 |
